# Factors associated with excess skin after bariatric surgery: A mixed method study

**DOI:** 10.1101/2023.04.26.23289160

**Authors:** Aurélie Baillot, Jennifer Brunet, Lucie Lemelin, Shaina A. Gabriel, Marie-France Langlois, André Tchernof, Laurent Biertho, Rémi Rabasa-Lhoret, Pierre Y. Garneau, Annie Aimé, Stéphane Bouchard, Ahmed J. Romain, Paquito Bernard

## Abstract

**Introduction:** After metabolic and bariatric surgery (MBS), many patients have excess skin (ES), which can cause inconveniences. Identifying factors related to ES quantity and inconveniences is crucial to inform interventions. The aim of this study was to identify sociodemographic, physical, psychosocial, and behavioral factors associated with ES quantity and inconveniences.

**Methods:** A mixed-method study with a sequential explanatory design was conducted with 124 adults (92% women, M_age_ 46.5±9.9 years, M_time post-MBS_ 34.2±27.6 months). During Phase I, ES quantity (arms, abdomen, thighs) and inconveniences, sociodemographic, anthropometric, clinical and behavioral outcomes were assessed. In Phase II, 7 focus groups were performed with 37 participants from Phase I. A triangulation protocol was completed to identify convergences, complementarities, and dissonances from quantitative and qualitative data.

**Results:** Quantitative data indicate only ES quantity on arms was associated with ES inconveniences on arms (r=.36, p<.01). Total ES quantity was associated with maximal body mass index (BMI) reached pre-MBS (r=.48, p<.05) and current BMI (r=.35, p<.05). Greater ES inconveniences was associated with higher social physique anxiety and age (R^2^=.50, p<.01). Qualitative data were summarized into 4 themes: psychosocial experiences living with ES, physical ailments due to ES, essential support and unmet needs, and beliefs of ES quantity causes.

**Conclusions:** Measured ES quantity is related to higher BMI, but not reported inconveniences. Greater self-reported ES quantity and inconveniences were associated with body image concerns.

## Introduction

Metabolic and bariatric surgery (MBS) is increasingly offered to improve health and quality of life in individuals with severe obesity[1]. However, significant weight loss post-MBS may result in excess skin (ES) in more than 70% of patients[2]. ES can be distressing and affect health and quality of life because of physical/medical (e.g., infections), psychosocial (e.g., shame, body dissatisfaction), and functional (e.g., hygiene, clothing) challenges[2]. Between 60 to 80% of patients want plastic surgery to remove ES post-MBS[3]; however, they are required to be weight stable for at least 6 months and access to such interventions remains limited. Consequently, patients have to contend with ES, at least temporarily.

ES quantity is variable; yet, factors to help explain observed interindividual differences in ES quantity remain elusive[2, 4, 5]. Sex/gender, but not age or time since surgery, relates to ES quantity[2, 4, 5], and results are equivocal for other factors (e.g., current BMI, pre-MBS BMI, amount of weight lost)[2, 4]. Likewise, the spectrum of inconveniences caused by ES patients report is variable. Putative risk factors include female sex/gender and body image concerns[2, 5–7]. Other factors such as depression, age, weight loss post-MBS, BMI prior to MBS, MBS type, time since MBS, and current BMI have been investigated, with conflicting or insufficient evidence[2]. Additional research is required to better understand factors associated with ES quantity and inconveniences to identify patients in greater need of intervention and inform intervention strategies.

ES measurement is at the forefront of investigating ES quantity and inconveniences. Self-reported ES (Sr-ES) quantity is common because it may be the only method available in large-scale survey studies. Although convenient, it is prone to bias and misreporting. As a result, methods relying on measured ES (M-ES) quantity are employed. Unfortunately, because few studies have investigated these outcomes together, evidence is lacking to ascertain if Sr-ES and M-ES quantity are associated (and provide evidence of convergent validity), and whether they relate to inconveniences caused by ES[4, 8].

### Objectives

A mixed-methods study was employed to combine quantitative and qualitative data to offer an in-depth understanding of ES experiences. Previous studies[2] and theoretical perspectives guided the choice of potential factors associated with ES quantity and inconveniences to investigate (details supplemental File 1). Quantitative objectives were: i) describe the relationships between M-ES quantity, Sr-ES quantity, and inconveniences caused by ES in adults post-MBS, and ii) assess the relationships of M-ES and Sr-ES quantity and inconveniences caused by ES with sociodemographic, physical, psychosocial, and behavioral factors. Qualitative objectives were: i) support (or refute) and supplement quantitative results by exploring factors related to ES quantity and inconveniences in adults post-MBS, and ii) expand quantitative results by exploring how adults’ ES quantity and inconveniences evolve over time, and patients’ support needs regarding ES management. Finally, the mixed objective was to integrate results on factors associated with ES quantity and inconveniences caused by ES to garner a better understanding of ES-related experiences in adults post-MBS.

## Materials and Methods

### Design

This is a mixed methods study with a sequential explanatory design (details supplemental File 2). The reporting herein complies with the Mixed Methods Article Reporting Standards (MMARS) and the Reporting Involvement of Patients and the Public (GRIPP2 short form). Human research ethics committees approved the study protocol (details supplemental File 2), and participants provided informed consent prior to data collection. Study objectives and different elements of the research process were informed by patient partners (details supplemental File 2).

### Sampling, recruitment, and data collection procedures

Sampling, recruitment, and data collection occurred in two phases. Recruitment for Phase I (quantitative phase) occurred from February 2019 to October 2021 via Facebook^©^, posters in waiting rooms of evaluation sites, and clinicians’ referral. By phone, a research assistant ensured participants met the following inclusion criteria: age ≥18 years, French- or English-speaking, and having undergone MBS >18 months ago. Current pregnancy, plastic surgery to remove ES, and >1 MBS were exclusion criteria. Eligible patients took part in an in-person 2-hour visit to have anthropometric measurements and pictures taken by a research assistant. They also completed an online survey via LimeSurvey^©^. Participants received $25 CAD. Supplemental File 2 presents the evaluation sequence.

Phase II (qualitative phase) consisted of focus groups with questions developed based on the results of a scoping review[2] and preliminary analysis of Phase I data (supplemental File 2).

Consenting Phase I participants who had ES were invited to participate in a focus group. Seven focus groups were conducted via Zoom©; 6 for women (June 2020 to March 2021), and 1 for men (January 2022). Focus groups were facilitated by LL, co-led by AB, and attended by a research assistant who took notes to summarize content. Focus groups comprised 3-8 participants, lasted 67-98 minutes, were video and audio recorded, and transcribed into verbatim by CV and AD. As all participants spoke French, all focus groups were conducted in French.

### Sample size

For Phase I, a preliminary analysis using nQuery Advisor software^©^ showed 124 participants were required to detect statistically significant correlations of ≥.25 with 80% power. For Phase II, to balance representativeness and saturation goals, all consenting Phase I participants who had ES and were available were included. This yielded 7 focus groups, which was considered appropriate as 3-6 groups typically reach data saturation[9].

### Quantitative Measures

<u>M-ES</u> quantity was measured on the dominant side of the abdomen, upper arms, and inner thighs using standardized photographs taken by 4 research assistants, one in each recruiting center. Research assistants indicated fixed anatomical structures before the photographs were taken using a marker to have reference points. Independently, evaluators based measurements on the adapted Iglésias scale[10]. Measurements were recorded as percentages. Evaluators averaged scores were compared, and percentage discrepancies exceeding 3% were discussed to reach consensus. Then, percentages for abdomen, upper arms, and inner thighs were averaged to obtain total M-ES[10]. <u>Sr-ES</u> quantity on the abdomen, upper arms, and inner thighs were collected using the Sahlgrenska Excess Skin (SES) questionnaire[11]. Total Sr-ES quantity was calculated by averaging responses. <u>Inconveniences caused by ES</u> were assessed using the SES[11] subscales for the abdomen, upper arms, inner thighs, and entire (total) body.

Participants had their height and body mass measured using a height rod and an electronic scale to calculate current BMI [weight (kg) / height (m)^2^]. Additionally, participants were asked to complete an online survey (Limesurvey^©^) to assess <u>sociodemographics</u> (age, sex, ethnicity, civil status, employment status, income level, highest level of education attained, perceived socioeconomic status)[12]. The survey was also used to collected <u>self-reported anthropometrics and weight history</u> (details supplemental File 2), and <u>clinical data</u> (number of past pregnancy(ies), skin illness(es), health conditions [hypertension, type 2 diabetes, sleep apnea, heart diseases, asthma, arthritis and/or arthrosis], MBS type, place, and date, striae quantity on the abdomen, thighs/buttocks, and hips [1 “no”, 2 “less than five”, 3 “ 5 to 10”, 4 “or more than 10 striae”][13], the presence of skin irritation or fungal infection because of ES [yes/no]. A total score for striae was computed by summing scores for abdomen, thighs/buttocks, and hips, ranging from 4 to 12. Finally, the survey comprised the following measures to collect data on psychosocial factors and lifestyle behaviors (details supplemental File 2): body dissatisfaction [*pictorial BMI-based body size guides* (BSGs)[14]], body esteem (appearance, attitude, weight) [*Body Esteem Scale for Adolescents and Adults* (BESAA)[15]], awareness and internalization of sociocultural body image norms [*Sociocultural Attitudes Towards Appearance Questionnaire 4 (SATAQ4)*[16]], social physique anxiety [*Social Physique Anxiety Scale (SPAS)*[17]], anxiety and depression symptoms [*Hospital Anxiety and Depression Scale (HADS)*[18]], perceived social support (satisfaction and availability) [*Bruchon-Schweitzer Scale (SSQ6)*[19]], marital satisfaction [*Dyadic Adjustment Scale (DAS-4*)[20]], quality of life [*Quality of Life, Obesity, and Dietetics (QOLOD) rating scale*[21]], current smoking status, alcohol [*Alcohol Use Disorder Identification Test-Concise* (AUDIT-C)[22]], sun exposure and sunscreen use, moderate-to-vigorous intensity physical activity and sedentary time [*Global physical activity questionnaire* (GPAQ)[23]].

### Quantitative Data Analysis

Preliminary analyses performed are described in supplemental File 2. To describe the relationships between M-ES quantity, Sr-ES quantity, and inconveniences caused by ES, bivariate correlations were performed. To examine the associations of ES quantity (Sr- and M) and inconveniences caused by ES with sociodemographic, physical, psychosocial, and behavioral factors, multivariate analyses were performed. Specifically, owing to the nested data (assessment sites) and missing data, mixed models were estimated. Bivariate correlations were estimated to include in the multivariate models only independent variables associated at p<.20. Two models were tested: one for Sr-ES quantity and one for inconveniences caused by ES. No model was tested for M-ES quantity because only maximal BMI reached and measured BMI were associated with it in bivariate analyses. Statistical analyses were carried out using R and the nlme and ggstatplot packages. Ps<.05 were interpreted as statistically significant.

### Qualitative Data Analysis

Focus group transcripts were imported into NVivo^©^12 and analyzed by a multidisciplinary team (LL, SG, AB, and JB; supplemental File 2 for details). The analysis was inspired by hybrid deductive and inductive thematic analysis approach[24], and steps followed are described in supplemental File 2. Data saturation was achieved during data analysis, whereby *a priori codes* were adequately represented in the data and ‘new’ codes stopped appearing during the analysis.

### Data Integration

Quantitative and qualitative data were integrated at the method level with the use of qualitative data collection to (dis)corroborate, supplement, and extend quantitative results, and at the interpretative and reporting level through narrative analysis and table comparison[25]. For the latter, a triangulation protocol was completed by LL, SG, AB, and JB to combine quantitative and qualitative findings into a table to assess if results from both methods were convergent (i.e. offer similar insights), complementary (i.e., enhance understanding), or dissonant (i.e. offer different views or lack of supportive narrative)[26].

## Results

### Quantitative Findings

Initially, 344 people expressed interested in this study (43% self-referred from Facebook^©^ advertisements, 26 % referred by clinicians, 31 % from unknown sources). Of these, 94 (27%) were not eligible because they underwent MBS <18 months ago (n=53; 56%), were currently pregnant (n=1; 1%), had plastic surgery to remove ES (n=24; 26%), and had >1 MBS (n=16; 17%). Moreover, 76 (22%) declined to participate and 50 (15%) could not be reached. In total, 124 (36%) provided informed consent and were enrolled. Participant characteristics for the total sample and by assessment sites are presented in Table 1 and supplemental File 3, respectively.

**Table 1.** Characteristics of the participants and bivariate correlations with ES quantity, and inconveniences caused by ES

| Outcomes | N | Mean±SD or % | Correlations with Total ES quantity: r | Correlations with total self-reported ES quantity: r | Correlations with total inconveniences caused by ES: r |
| --- | --- | --- | --- | --- | --- |
| **ES outcomes** |  |  |  |  |  |
| Total ES quantity (%) | 112 | 60.5±20.4 | / | 0.41* | 0.08 |
| Abdomen ES quantity (%) | 112 | 81.5±27.4 | 0.72* | 0.33* | 0.08 |
| Arms ES quantity (%) | 124 | 68.3±35.0 | 0.88* | 0.35* | 0.14 |
| Thighs ES quantity (%) | 124 | 30.1±17.2 | 0.59* | 0.26 | -0.03 |
| Total self-reported ES score | 115 | 2.6±0.8 | 0.41* | / | 0.59* |
| Abdomen self-reported ES score | 115 | 2.8±0.9 | 0.27 | 0.77* | 0.50* |
| Arms self-reported ES score | 116 | 2.4±0.9 | 0.48* | 0.75* | 0.42* |
| Thighs In self-reported ES score | 115 | 2.5±1.1 | 0.25 | 0.85* | 0.49* |
| Total inconveniences caused by ES score | 124 | 7.2±2.4 | 0.08 | 0.59* | / |
| Abdomen inconveniences caused by ES score | 123 | 7.4±2.7 | 0.09 | 0.57* | 0.81* |
| Arms inconveniences caused by ES score | 121 | 6.5±2.8 | 0.20 | 0.59* | 0.69* |
| Thighs In inconveniences caused by ES score | 122 | 6.8±2.9 | 0.19 | 0.70* | 0.69* |
| **Sociodemographic data** |  |  |  |  |  |
| Sex (% of women) | 124 | 91.9 | NP | NP | NP |
| Age (years) | 124 | 46.5±9.9 | 0.08 | 0.06 | 0.13 |
| Ethnicity (% White) | 124 | 88.7 | 0.11 | 0.11 | 0.03 |
| Civil status (% married or common law partner) | 122 | 60.7 | -0.07 | -0.08 | 0.02 |
| Perceived socioeconomic status score | 124 | 5.4±1.7 | -0.02 | 0.21 | 0.28 |
| Full-time worker (%) | 124 | 75.8 | 0.13 | 0.15 | 0.17 |
| **Anthropometric data and weight history** |  |  |  |  |  |
| Body mass index (kg.m<sup>2</sup>) | 124 | 31.0±6.0 | 0.35* | 0.13 | -0.12 |
| Period of maximal weight: | 124 |  | 0.02 | -0.18 | -0.09 |
| - adolescence/young adulthood | 26 | 21.0% | / | / | / |
| - adulthood (30-50 years old) | 83 | 67.0% | / | / | / |
| - after 50 years old | 15 | 12.1% | / | / | / |
| Maximal body mass index reached (kg.m<sup>2</sup>) | 124 | 51.9±9.5 | 0.40* | 0.24 | -0.15 |
| Weight loss (kg) | 123 | 57.4±21.3 | 0.02 | 0.07 | -0.18 |
| **Clinical data** |  |  |  |  |  |
| MBS type | 124 |  | 0.28 | 0.05 | 0.07 |
| - Biliopancreatic diversion | 28 | 22.6% | / | / | / |
| - Sleeve gastronomy | 59 | 47.6% | / | / | / |
| - Gastric bypass | 30 | 24.2% | / | / | / |
| - Other | 7 | 5.6% | / | / | / |
| Time since MBS (months) | 123 | 34.2±27.6 | 0.03 | 0.04 | -0.01 |
| Total stria number score | 124 | 8.5±2.8 | 0.09 | 0.25 | 0.19 |
| Presence of skin fungal infection because of ES | 54 | 43.5% | 0.01 | 0.22 | 0.21 |
| Presence of skin irritation because of ES | 81 | 65.3% | 0.01 | 0.22 | 0.21 |
| Presence of type 2 diabetes | 2 | 1.6% | 0.06 | 0.21 | 0.15 |
| Presence of hypertension | 15 | 12.2% | -0.09 | -0.11 | 0.01 |
| Presence of heart diseases | 4 | 3.3% | 0.01 | 0.1 | -0.01 |
| Presence of sleep apnoea | 15 | 12.2% | 0.11 | -0.09 | -0.25 |
| Presence of asthma | 16 | 13.0% | 0.03 | 0.22 | 0.09 |
| Presence of arthritis and/or arthrosis | 17 | 13.8% | 0.07 | 0.15 | 0.1 |
| Presence of eczema | 12 | 9.7% | -0.01 | -0.14 | -0.15 |
| Presence of previous pregnancy | 84 | 73.7% | -0.13 | -0.02 | 0.06 |
| **Psychosocial data** |  |  |  |  |  |
| Body dissatisfaction | 124 | 2.4±1.8 | 0.23 | 0.36* | 0.17 |
| Body-esteem appearance | 124 | 1.6±0.8 | 0 | -0.53* | -0.57* |
| Body-esteem attitude | 124 | 2.1±0.6 | -0.06 | -0.14 | -0.09 |
| Body-esteem weight | 124 | 1.8±0.8 | -0.06 | -0.3* | -0.32* |
| Internalization of sociocultural body image | 124 | 57.0±17.8 | -0.04 | 0.27 | 0.33* |
| Social physique anxiety | 124 | 32.5±8 | -0.08 | 0.51* | 0.65* |
| Social support satisfaction | 124 | 31.3±5.1 | 0.07 | -0.05 | -0.12 |
| Social support availability | 124 | 18.9±9.4 | 0.11 | -0.07 | -0.25 |
| Marital satisfaction | 87 | 15.9±3.8 | 0.08 | -0.06 | -0.1 |
| Anxiety symptoms | 124 | 8.1±4 | -0.04 | 0.21 | 0.31* |
| Depression symptoms | 124 | 3.4±3.2 | -0.07 | 0.14 | 0.12 |
| Quality of life | 124 | 61.4±15.5 | -0.03 | -0.29* | -0.32* |
| **Behavioral data** |  |  |  |  |  |
| Sedentary time (h/d) | 124 | 8.0±3.5 | -0.07 | 0.01 | -0.09 |
| Moderate-to-vigorous intensity physical activity (min/wk) | 124 | 188.4±251.3 | -0.01 | -0.18 | 0.1 |
| Non-smoker (%) | 124 | 91.1% | -0.03 | -0.11 | -0.09 |
| Alcohol AUDIT C score | 124 | 4.7±2.6 | -0.03 | -0.11 | -0.08 |
| Sun exposure | 124 | 2.4±0.8 | -0.13 | 0.05 | 0.07 |

\* = $p \leq .05$ ; AUDIT = Alcohol Use Disorders Identification Test; ES= excess skin; MBS=metabolic and bariatric surgery; NP=not performed because of the small sample of men; SD=standard deviation.

### Associations between ES quantity and inconveniences

Figure 1 illustrates bivariate correlations between total ES quantity variables and total inconveniences caused by ES. Supplemental File 4 shows bivariate correlations between ES outcomes (total, abdomen, arms, and thighs). M-ES quantity was not significantly associated with inconveniences caused by ES (total, abdomen, and inner thighs, ps>.05), except for arms (r=.36; p=.002). M-ES quantity and Sr-ES quantity were significantly correlated for total (r=.41, p<.001), arms (r=.52, p <.001), and abdomen (r=.43; p<.001), but not for inner thighs (r=.28, p=.06). Sr-ES quantity was significantly associated with inconveniences caused by ES for total (r=.59, p<.001), abdomen (r=.63, p<.001), arms (r=.71, p<.001), and inner thighs (r=.76; p<.001).

**Figure 1.**
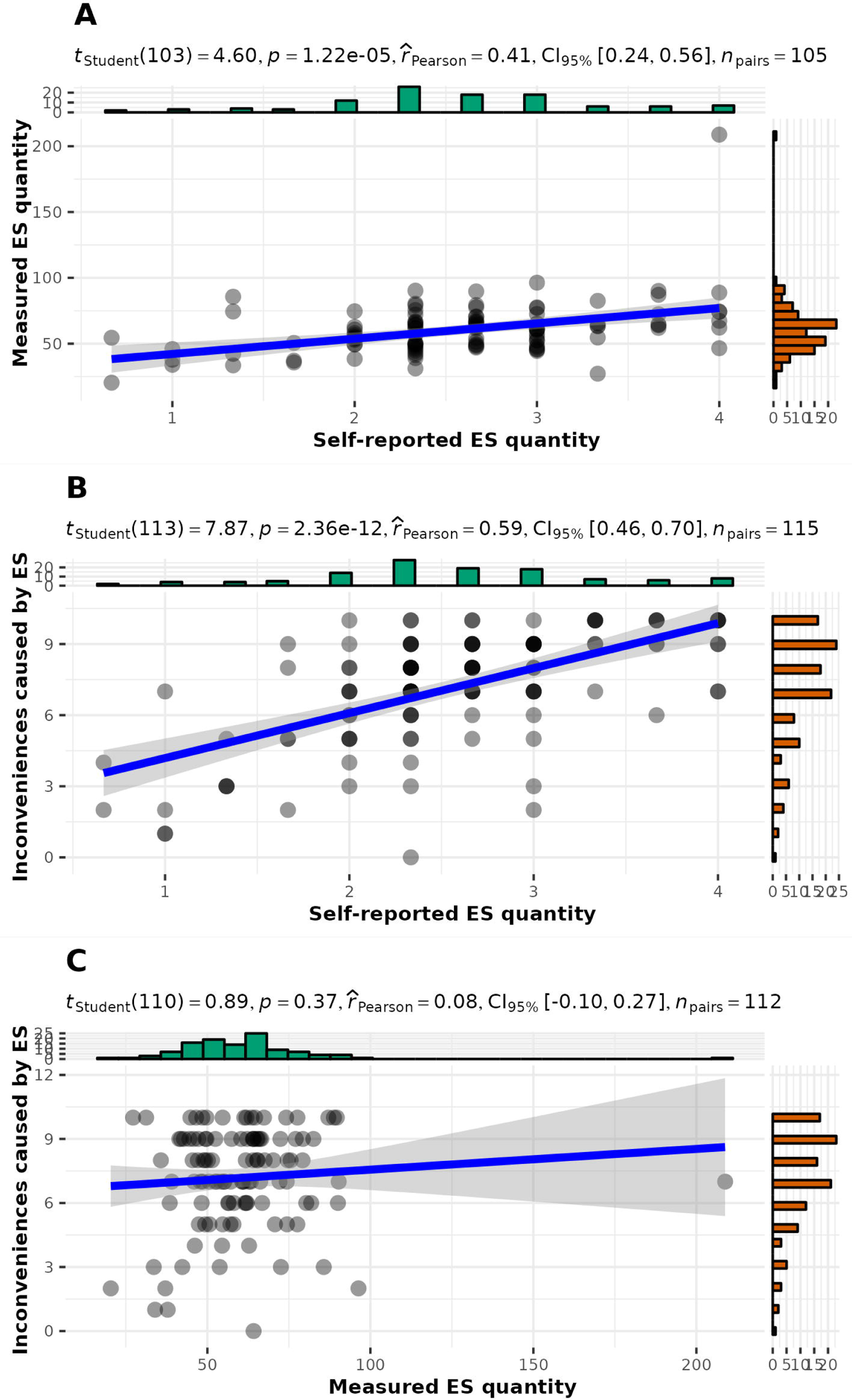
Bivariate correlations between total ES quantity and total self-reported ES quantity (A), total self-reported ES quantity and total inconveniences caused by ES (B), total ES quantity and total inconveniences caused by ES (C) in participants at least 18 months after MBS.

### Associations of ES quantity and inconvenience with other factors

Table 1 presents bivariate correlations between total ES quantity variables, total inconveniences caused by ES, and sociodemographic, physical, psychosocial, and behavioral factors (see supplemental File 5 for details results with ES on abdomen, arms, and thighs). For total Sr-ES quantity, multivariate analyses revealed positive associations with social physique anxiety, body dissatisfaction and age, and a negative association with body esteem (R^2^=.38, p<.05) (Table 2). For total inconveniences caused by ES, multivariate analyses revealed positive association with social physique anxiety and age (R^2^=.50, p<.01) (Table 2).

**Table 2.**
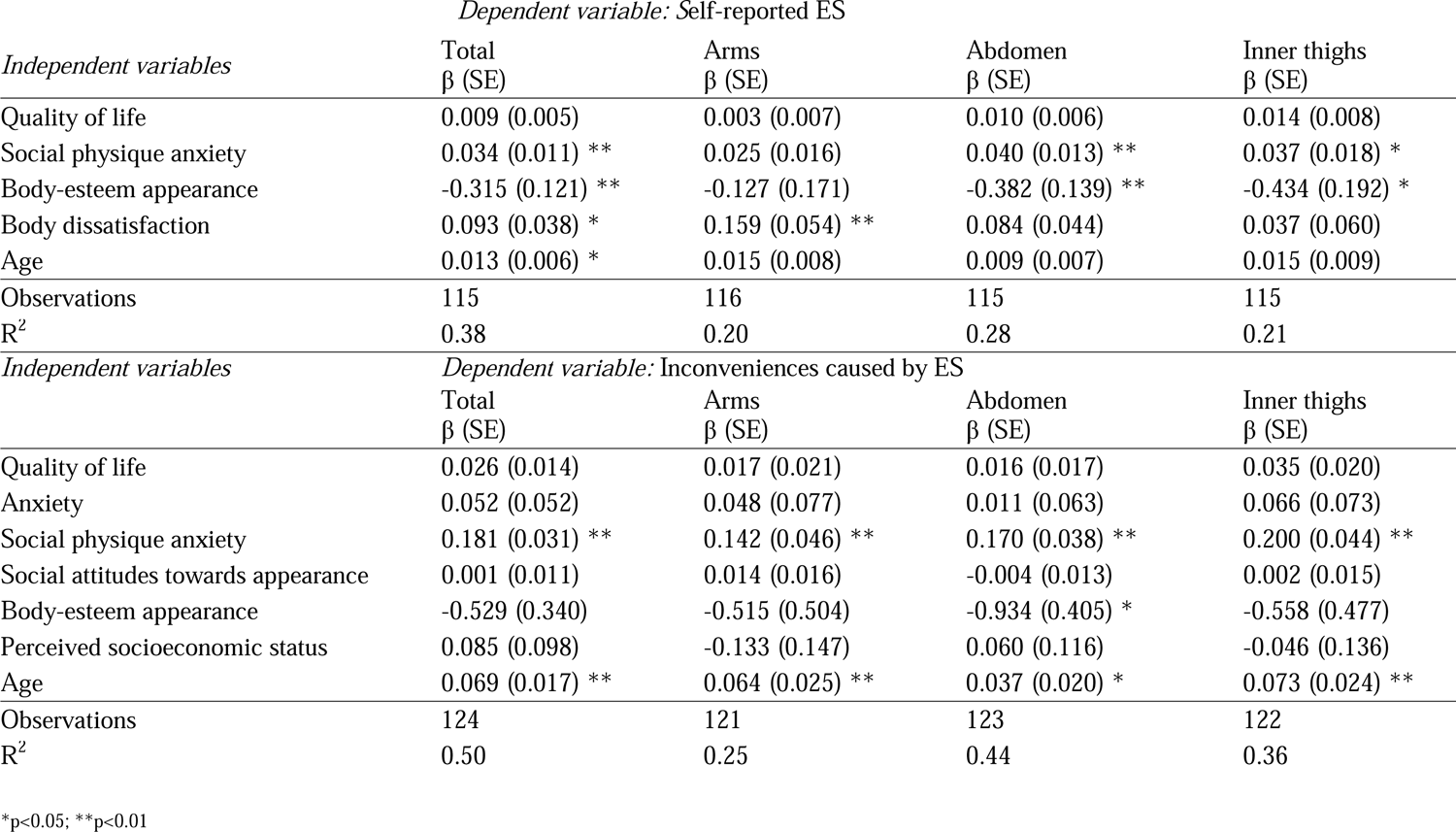
Multivariate analyses to explain self-reported ES quantity and inconveniences caused by ES in participants after metabolic and bariatric surgery

| *Independent variables* | *Dependent variable: Self-reported ES* |  |  |  |
| --- | --- | --- | --- | --- |
| | Total<br>$\beta$ (SE) | Arms<br>$\beta$ (SE) | Abdomen<br>$\beta$ (SE) | Inner thighs<br>$\beta$ (SE) |
| Quality of life | 0.009 (0.005) | 0.003 (0.007) | 0.010 (0.006) | 0.014 (0.008) |
| Social physique anxiety | 0.034 (0.011) ** | 0.025 (0.016) | 0.040 (0.013) ** | 0.037 (0.018) * |
| Body-esteem appearance | -0.315 (0.121) ** | -0.127 (0.171) | -0.382 (0.139) ** | -0.434 (0.192) * |
| Body dissatisfaction | 0.093 (0.038) * | 0.159 (0.054) ** | 0.084 (0.044) | 0.037 (0.060) |
| Age | 0.013 (0.006) * | 0.015 (0.008) | 0.009 (0.007) | 0.015 (0.009) |
| Observations | 115 | 116 | 115 | 115 |
| R<sup>2</sup> | 0.38 | 0.20 | 0.28 | 0.21 |
| *Independent variables* | *Dependent variable: Inconveniences caused by ES* |  |  |  |
| | Total<br>$\beta$ (SE) | Arms<br>$\beta$ (SE) | Abdomen<br>$\beta$ (SE) | Inner thighs<br>$\beta$ (SE) |
| Quality of life | 0.026 (0.014) | 0.017 (0.021) | 0.016 (0.017) | 0.035 (0.020) |
| Anxiety | 0.052 (0.052) | 0.048 (0.077) | 0.011 (0.063) | 0.066 (0.073) |
| Social physique anxiety | 0.181 (0.031) ** | 0.142 (0.046) ** | 0.170 (0.038) ** | 0.200 (0.044) ** |
| Social attitudes towards appearance | 0.001 (0.011) | 0.014 (0.016) | -0.004 (0.013) | 0.002 (0.015) |
| Body-esteem appearance | -0.529 (0.340) | -0.515 (0.504) | -0.934 (0.405) * | -0.558 (0.477) |
| Perceived socioeconomic status | 0.085 (0.098) | -0.133 (0.147) | 0.060 (0.116) | -0.046 (0.136) |
| Age | 0.069 (0.017) ** | 0.064 (0.025) ** | 0.037 (0.020) * | 0.073 (0.024) ** |
| Observations | 124 | 121 | 123 | 122 |
| R<sup>2</sup> | 0.50 | 0.25 | 0.44 | 0.36 |

\*p<0.05; \*\*p<0.01

### Qualitative Findings

Of the 124 Phase I participants, 37 (30%) took part in a focus group. Of the remaining, 34 (27%) declined to take part in the focus groups, 45 (36%) did not return our calls, 8 (6%) were unavailable for the focus groups. Focus group participant characteristics are presented Table 3. Focus group data were summarized into 4 main themes with several subthemes (Table 4 and supplemental File 6).

**Table 3.**
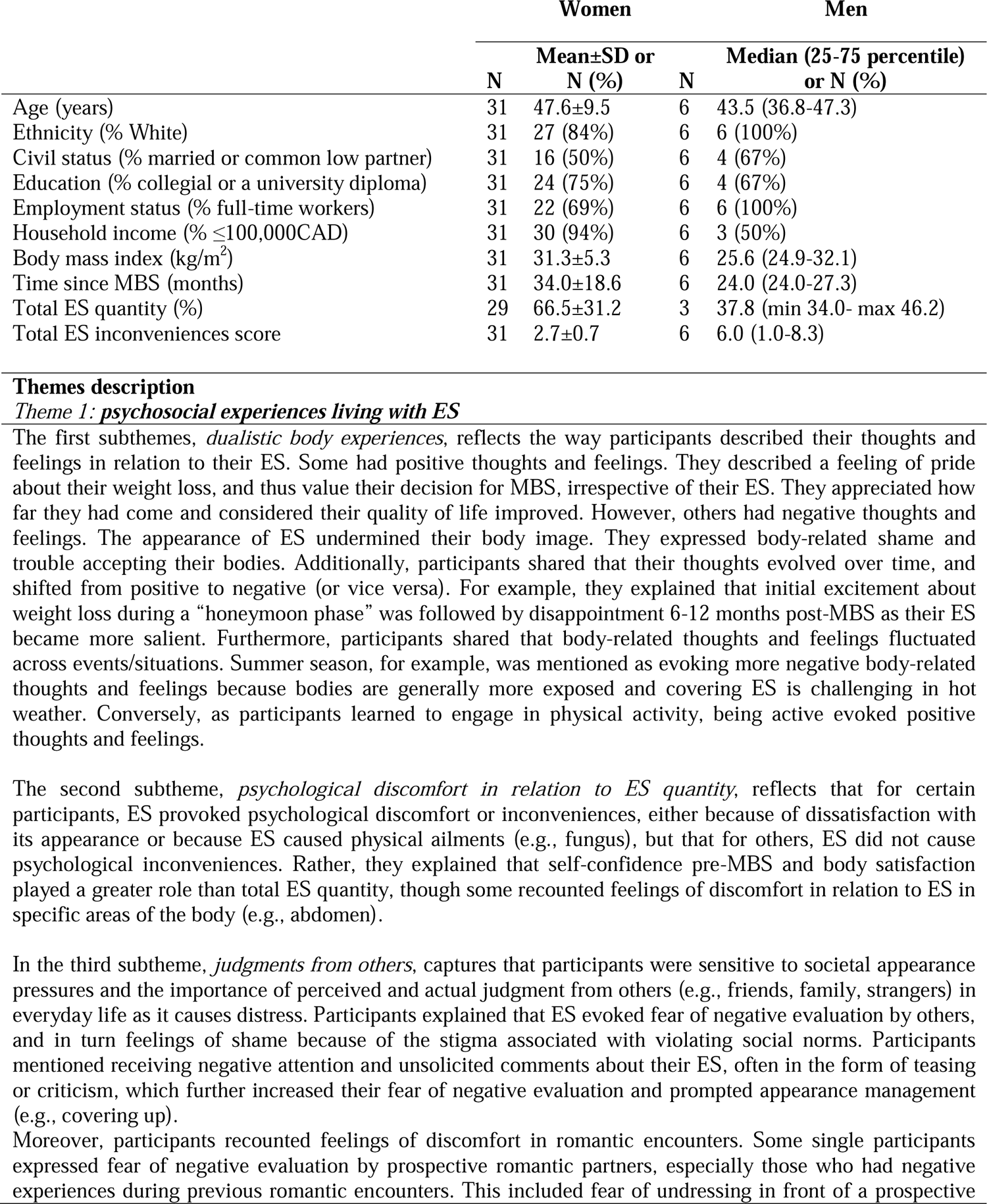
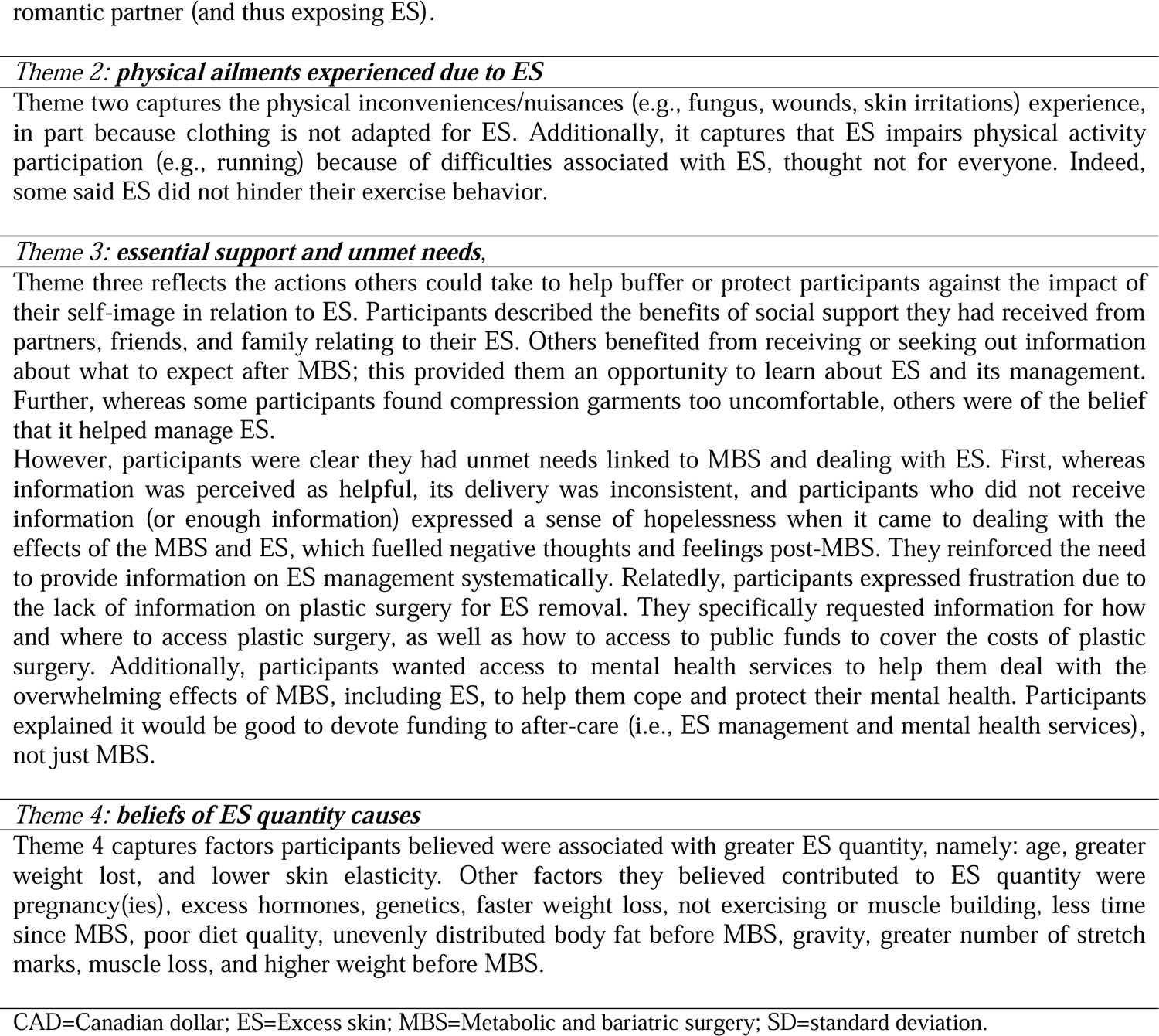
Participant characteristics of the focus group and results

|  | Women |  | Men |  |
| --- | --- | --- | --- | --- |
|  | N | Mean±SD or N (%) | N | Median (25-75 percentile) or N (%) |
| Age (years) | 31 | 47.6±9.5 | 6 | 43.5 (36.8-47.3) |
| Ethnicity (% White) | 31 | 27 (84%) | 6 | 6 (100%) |
| Civil status (% married or common law partner) | 31 | 16 (50%) | 6 | 4 (67%) |
| Education (% collegial or a university diploma) | 31 | 24 (75%) | 6 | 4 (67%) |
| Employment status (% full-time workers) | 31 | 22 (69%) | 6 | 6 (100%) |
| Household income (% ≤100,000CAD) | 31 | 30 (94%) | 6 | 3 (50%) |
| Body mass index (kg/m<sup>2</sup>) | 31 | 31.3±5.3 | 6 | 25.6 (24.9-32.1) |
| Time since MBS (months) | 31 | 34.0±18.6 | 6 | 24.0 (24.0-27.3) |
| Total ES quantity (%) | 29 | 66.5±31.2 | 3 | 37.8 (min 34.0- max 46.2) |
| Total ES inconveniences score | 31 | 2.7±0.7 | 6 | 6.0 (1.0-8.3) |

**Themes description*****Theme 1: psychosocial experiences living with ES***
The first subthemes, *dualistic body experiences*, reflects the way participants described their thoughts and feelings in relation to their ES. Some had positive thoughts and feelings. They described a feeling of pride about their weight loss, and thus value their decision for MBS, irrespective of their ES. They appreciated how far they had come and considered their quality of life improved. However, others had negative thoughts and feelings. The appearance of ES undermined their body image. They expressed body-related shame and trouble accepting their bodies. Additionally, participants shared that their thoughts evolved over time, and shifted from positive to negative (or vice versa). For example, they explained that initial excitement about weight loss during a “honeymoon phase” was followed by disappointment 6-12 months post-MBS as their ES became more salient. Furthermore, participants shared that body-related thoughts and feelings fluctuated across events/situations. Summer season, for example, was mentioned as evoking more negative body-related thoughts and feelings because bodies are generally more exposed and covering ES is challenging in hot weather. Conversely, as participants learned to engage in physical activity, being active evoked positive thoughts and feelings.
The second subtheme, *psychological discomfort in relation to ES quantity*, reflects that for certain participants, ES provoked psychological discomfort or inconveniences, either because of dissatisfaction with its appearance or because ES caused physical ailments (e.g., fungus), but that for others, ES did not cause psychological inconveniences. Rather, they explained that self-confidence pre-MBS and body satisfaction played a greater role than total ES quantity, though some recounted feelings of discomfort in relation to ES in specific areas of the body (e.g., abdomen).
In the third subtheme, *judgments from others*, captures that participants were sensitive to societal appearance pressures and the importance of perceived and actual judgment from others (e.g., friends, family, strangers) in everyday life as it causes distress. Participants explained that ES evoked fear of negative evaluation by others, and in turn feelings of shame because of the stigma associated with violating social norms. Participants mentioned receiving negative attention and unsolicited comments about their ES, often in the form of teasing or criticism, which further increased their fear of negative evaluation and prompted appearance management (e.g., covering up).
Moreover, participants recounted feelings of discomfort in romantic encounters. Some single participants expressed fear of negative evaluation by prospective romantic partners, especially those who had negative experiences during previous romantic encounters. This included fear of undressing in front of a prospective
romantic partner (and thus exposing ES).
***Theme 2: physical ailments experienced due to ES***
Theme two captures the physical inconveniences/nuisances (e.g., fungus, wounds, skin irritations) experience, in part because clothing is not adapted for ES. Additionally, it captures that ES impairs physical activity participation (e.g., running) because of difficulties associated with ES, though not for everyone. Indeed, some said ES did not hinder their exercise behavior.
***Theme 3: essential support and unmet needs,***

### Mixed Method Findings (Integration)

Supplemental File 7 presents wherein quantitative and qualitative findings were convergent, complementary, or dissonant.

## Discussion

The purpose of this study was to explore and describe the relationships between M-ES quantity, Sr-ES quantity and inconveniences caused by ES, and to extend knowledge on factors associated with ES quantity and inconveniences. According to quantitative results, M-ES quantity is not systematically related to inconveniences caused by ES, whereas Sr-ES quantity is positively correlated with inconveniences caused by ES. These results are consistent with other studies reporting an association between Sr-ES quantity and inconveniences[27, 28]. However, results are discordant between studies when it comes to associations between inconveniences caused by ES and M-ES quantity[2, 8, 29]. This discrepancy is well illustrated by participants’ conflicting views within our qualitative findings (i.e., whereby some participants, but not others, believed greater ES quantity leads to greater inconveniences caused by ES). Beyond individuals’ holding different perspectives, such inconsistent results could be explained by the methods used to assess ES (e.g., directly or photo), the different population studied (e.g., gender, country), or the timing of assessments (e.g., time elapse after surgery, season). Also, the quantitative and qualitative findings suggest it is desirable to distinguish between location of ES, as location played a role in the experience of inconveniences caused by ES and the associations observed within the quantitative data were influenced by the location of ES (arms vs thigh and abdomen). Last, it is possible that visibility and/or interference with mouvements could explain this observation.

In addition, the quantitative results show Sr-ES quantity is positively associated with M-ES quantity, as reported in two others studies[8, 28]. However, ES inconvenience was related to Sr-ES quantity, but not systematically with M-ES quantity. This suggests having participants estimate their own ES quantity may be, in part, influenced by their experiences of inconveniences. Arguably, individual with more inconveniences caused by ES could overestimate their ES quantity. Additional studies should assess the validity of Sr-ES quantity and identify factors associated with under/over-estimation. Results also emphasize the value of considering both because yield valuable data (i.e., individuals’s subjective accounts of their ES for Sr-ES and diagnositic information for M-ES). Moreover, this could provide new opportunity to study intrapersonal and interpersonal variability in these constructs.

A better understanding of factors associated with ES quantity post-MBS is essential to help identify patient pre-MBS at risk of greater ES, and prepare them to realistic expectations[8]. In line with prior studies[4, 8, 29], quantitative and qualitative results suggest maximal BMI, pre-MBS and current BMI, but not psychosocial factors, are related to ES quantity post-MBS. As these findings were retrospective, these results need to be confirmed in prospective longitudinal studies.

Interestingly, qualitative results suggest skin elasticity and quality, and previous pregnancy(ies) can lead to more ES quantity post-MBS. While not supported by quantitative results and previous studies[8], past pregnancy seems to be a reason often given probably because of its impact on skin elasticity. Although stretch marks were explored herein as a proxy of skin quality, no association was found with ES quantity. Additional studies with better indicators of skin quality and elasticity should be explored. In addition, qualitative findings call for investigating other variables related to ES quantity (e.g., diet quality and nutritional deficiencies, muscle mass, hormones, genetic).

Regarding inconveniences caused by ES, quantitative and qualitative findings both aligned with past studies[5–7] and offered similar insights; that, it is associated with body image factors such as social physique anxiety and body esteem. While there is evidence that MBS can improve some dimensions of body image, it is a complex and multidimensional concept, and accordingly not all dimensions may improve post-MBS[30], especially for individuals experiencing greater inconveniences caused by ES. Because negative body image factors relate to depressive symptoms and eating disorders, both associated with weight regain in past studies[6, 7] and alluted to by focus group participants, mental health support is needed to deal with important and rapid body changes post-MBS.

In this study, qualitative findings added new and crucial insights to the quantitative results. Notably, participants explained inconveniences caused by ES can evolve over time (e.g., after the “honeymoon phase” of weight loss) and change when ES is more exposed in certain events (e.g., seeing photos of oneself), situations (e.g., during intimacy and physical examinations by healthcare providers) and seasons (e.g., during summer, short sleeves, swimsuit). These potential variations support the clinical and scientific importance for repeated measurements, the addition of contextual information when assessing inconveniences caused by ES, and the delivery of support at various timepoints post-MBS to help patients.

Moreover, qualitative findings provided valuable insight into peoples’ perspectives related to the ES inconveniences-social support association. Indeed, they suggest ‘social support’ can be either positive or negative, and that if both are not considered, quantitative analyses may fail to reveal an association between inconveniences caused by ES and social support.

Regarding moderate-to-vigorous-intensity physical activity, quantitative results showed that ES quantity and inconveniences were not significantly related. However, qualitative results and those of Olsen et al.[31] bring relevant nuance to the absence of association. Indeed, participants seem to avoid certain types of physical activity (e.g., jumping, running) because of the weight of their ES. Thus, ES quantity may be a barrier to engaging in certain types of physical activity, thus requiring consideration when developing programs for this population. In addition, qualitative findings suggest a need to shift away from the current way of analyzing physical activity quantitatively (e.g., frequency, volume) and describe/analyze important elements of physical activity (i.e., type, setting (e.g., gym center or at home), ES exposition (e.g., swimsuit)) in relation to ES inconveniences.

### Methodological considerations

This study adds important knowledge to the current understanding of ES experiences post-MBS. Using complementary quantitative and qualitatives approaches ensured adults were given a voice to share their experiences and perspectives on what is associated with ES quantity and inconveniences. However, the study presents some limitations. Participants recruited were primarily female, and as such, their perspectives may not coincide with males’ perspectives. Moreover, recruitment of participants occurred in the province of Quebec (Canada) and mostly via social media. Thus, findings may not be representative of other populations. Additionally, the cross-sectional design precludes causal inferences. Moreover, several questionnaires not specific to adults who underwent MBS (e.g., social support, marital satisfaction), could have reduced construct validity. Finally, the researchers’ interests and backgrounds are recognized as potentially influencing the collection, analysis and interpretation of the qualitative data.

## Conclusion

This mixed method study suggests that inconveniences caused by ES are not systematically related to M-ES quantity. Participants’ narratives supplemented this result by showing this association could be influenced by the location of ES and situations where ES is exposed. Higher maximal BMI pre-MBS is associated with greater ES quantity post-MBS. The associations found between body image factors and ES inconveniences post-MBS were supported by participants’ narratives, emphasizing the need of support to manage body image to improve patients’ MBS experiences.

## Data Availability

All data produced in the present study are available upon reasonable request to the authors

## Acknowledgements

The authors would like to thank, Anne Bastin, Élise Bolteau, Christine Brown, Alexia Duciaume, Josée Gagnon, Mélanie Nadeau, Melissa Pelletier, Mylène Simonneau, and Cassandra Voyer for assisting with the recruitment of participants and data collection. The authors would also like to thank, Maryse Deraiche, Stéphane Bergeron, and Christyne Simard for their involment as patient partner. SB is supported by a Tier I Canada Research Chair in clinical cyberpsychology at UQO. AB, and PB are recipients of salary awards from the Fonds de recherche du Québec-Santé (FRQ-S). JB is supported by a Tier II Canada Research Chair in Physical Activity Promotion for Cancer Prevention and Survivorship.

